# The landscape of childhood vaccine exemptions in the United States

**DOI:** 10.1101/2020.08.13.20174532

**Authors:** Casey M Zipfel, Romain Garnier, Madeline Kuney, Shweta Bansal

**Affiliations:** Department of Biology, Georgetown University, Washington DC, USA

## Abstract

Once-eliminated vaccine-preventable childhood diseases, such as measles, are resurging across the United States. Understanding the spatio-temporal trends in vaccine exemptions is crucial to targeting public health intervention to increase vaccine uptake and anticipating vulnerable populations as cases surge. However, prior available data on childhood disease vaccination is either on too rough a spatial scale for this spatially-heterogeneous issue, or is only available for small geographic regions, making general conclusions infeasible. Here, we have collated school vaccine exemption data across the United States and provide it at the county-level for all years available. We demonstrate the fine-scale spatial heterogeneity in vaccine exemption levels, and show that many counties may fall below the herd immunity threshold. We also show that vaccine exemptions increase over time in most states, and non-medical exemptions are highly prevalent where allowed. Our dataset also highlights the need for greater data sharing and standardized reporting across the United States.

## Background & Summary

Sufficient vaccine coverage to achieve herd immunity is the key to eliminating vaccine-preventable childhood diseases [1]. In the United States, the resurgence of once-eliminated diseases such as measles has reignited discussions over the role of vaccine hesitancy in diminishing herd immunity [2]. State-mandated school entry immunization requirements in the United States play an important role in achieving high vaccine coverage, but variations in vaccine exemption policies result in a patchwork of vaccine coverage across the country [3]. In all states, medical exemptions from immunizations can be obtained for conditions such as those resulting in immuno-suppresion or in cases of known adverse reaction to past immunizations. These exemptions have traditionally remained at levels low enough to not represent a threat to herd immunity, although recent dynamics such as that observed in California may change this assessment in the future [4, 5, 6]. Most other states offer religious exemptions, in which individuals cite personal religious beliefs that preclude vaccination, while others allow philosophical exemptions, which encompasses any personal belief against receiving a vaccination. The difficulty to obtain these exemptions for the parents varies widely from state to state [7], helping to create a heterogeneous landscape of immunization policies and of vaccination coverage.

It is essential that we understand the variations in vaccination at spatial scales that are relevant for the circulation of vaccine preventable diseases and to the implementation of public health policies. Such an understanding requires fine-scale geographic data on vaccination coverage and hesitancy. In the United States, information on a number of medical and non-medical exemptions is maintained and is accessible through the Center for Disease Control and Prevention (CDC) [8]. However, this spatial scale is largely inadequate as both exemptions and vaccine preventable childhood disease cases tend to cluster at smaller spatial scales [9]. Many prior studies have been performed at smaller scales, from the county to the school levels [10, 11,3, 12, 13]. However, the datasets used in these studies tend to be limited both in time and in terms of the spatial area they cover. Larger scale studies remain a challenge because there is no unified resource for data on childhood vaccination at more local spatial scales [14]. Each state is generally responsible for sharing exemptions data, and, when the data is shared at all, the formats vary widely between states as well as within states for different years. We aim to bridge that gap and facilitate further spatial studies of medical and non-medical exemptions by providing a standardized dataset of medical and non-medical exemptions at the county-scale encompassing multiple states and years in the continental United States.

## Methods

We collected data from all US states where school vaccine exemption information was freely available from the Department of Health website in any format. We were able to locate that data in 24 states (see Table 1 for a list of states included). Within these states, the number of years available varied relatively widely, between 19 years in California and a single year in 6 states. The most represented year in our dataset was 2017 (corresponding to school year 2017-2018). Because the dataset was compiled in June-July 2019, we note that it is possible that additional data for recent years may not be available, or that data may have become available in additional states not included in our dataset.

**Table 1:** Exemption data reporting varies widely across states. The state-level data reported by year is represented by an “X” in the year it is reported. California also reports data from 2000-2006, but is omitted for simplicity. Temporal data reporting is very inconsistent.

| State | 2006 | 2007 | 2008 | 2009 | 2010 | 2011 | 2012 | 2013 | 2014 | 2015 | 2016 | 2017 | 2018 |
| --- | --- | --- | --- | --- | --- | --- | --- | --- | --- | --- | --- | --- | --- |
| AL |  |  |  |  |  |  |  |  | X | X | X | X |  |
| AZ |  |  |  |  |  |  |  | X | X | X | X | X | X |
| CA | X | X | X | X | X | X | X | X | X | X | X | X | X |
| CO |  |  |  |  |  |  |  |  |  |  | X |  |  |
| CT |  |  |  |  |  |  | X | X | X | X | X | X |  |
| FL |  | X | X | X | X | X | X | X | X | X | X |  |  |
| IA | X |  | X | X | X | X | X | X | X | X | X | X |  |
| ID |  |  |  |  |  |  |  |  |  |  |  | X |  |
| IL |  | X | X | X | X | X |  |  |  |  |  |  |  |
| MA |  |  |  |  |  |  |  | X | X | X | X | X |  |
| ME |  |  |  |  |  |  |  |  | X | X | X | X | X |
| MI |  |  |  |  |  |  |  |  |  |  |  | X |  |
| MN |  |  |  |  |  |  | X | X | X | X | X | X |  |
| NJ |  |  |  |  |  |  |  | X | X | X | X | X |  |
| NY |  |  |  |  |  |  | X | X | X | X | X | X |  |
| OR |  |  |  |  |  |  |  |  | X | X | X | X | X |
| PA |  |  |  |  |  | X | X | X | X | X | X | X |  |
| SD |  |  |  |  |  |  |  |  | X | X | X | X |  |
| TN |  |  |  |  |  |  |  |  |  |  |  | X |  |
| TX |  |  |  |  |  | X | X | X | X | X | X | X |  |
| VA |  |  | X | X | X | X | X | X | X | X | X | X | X |
| VT |  |  |  |  |  |  |  |  |  |  |  | X |  |
| WA |  |  |  |  |  |  |  | X | X | X | X | X |  |

The data format varied widely between states, and exemptions were reported either as a number of exemptions or as a percentage of the enrolled students. We have elected to use number of students rather than percentages, and have transformed data as needed. For most states included in our dataset, the data are provided at the county level. In several states (Arizona, Colorado, Illinois, Maine, Michigan, South Dakota, Tennessee, Vermont, Oregon, and Washington), the data was provided at the school level, which we aggregated to the county.

Additional data processing was necessary in some cases. In Virginia, data was provided by school name, but county or city information was not included. We used a list of public and private schools to match school names with their respective county using fuzzy matching (with the ‘fuzzywuzzy’ Python package) with an 80% matching requirement. Our algorithm was unable to find a suitable match for between 3.8% and 6.8% of schools (depending on year), and these schools were not included in the final counts at the county level. Similarly, in Idaho, data at the school level included city information but county was not provided. We first matched city and county names, before aggregating the exemption data at the county level. Finally in New York state, exemptions were provided as percentages at the school level but enrollment information was not included. We obtained enrollment for public and private schools separately from the New York State Education Department, and used the school unique code to calculate exemption number from enrollment and exemption percentages. We then aggregated these numbers at the county level.

States reported data for exemptions based on varying definitions, so we selected data records based on data availability to make the data comparable cross states. We aimed to achieve parsimonious definitions of total medical exemptions (Figure 1A), total non-medical exemptions (Figure 1B), and total exemptions (Figure 1C), which includes both types of exemptions. We define medical exemptions as reported total medical exemptions. In Florida, permanent medical exemptions were reported separately from temporary medical exemptions, so permanent medical exemptions was chosen to represent total medical exemptions. To define total non-medical exemptions, we considered the state law regarding non-medical exemptions and the data availability. If the state reported total aggregated non-medical exemptions, that was selected as total non-medical exemptions. If the state reported only religious exemptions and only allows religious exemptions, that was selected as total non-medical exemptions. If the state reported only religious exemptions, but also allows philosophical exemptions, that was considered missing data. If the state allows philosophical exemptions and only reports philosophical exemptions, that was selected as total non-medical exemptions, as the state may not differentiate religious from philosophical. If the state allows philosophical exemptions and reports both religious and philosophical exemptions separately, these values were summed for total non-medical exemptions. To define total exemptions, if the state reported a total exemptions value, this value was used. If the state did not report a total exemptions value, but reported values for total medical exemptions and total non-medical exemptions, as defined above, these were summed for total exemptions. If the state was missing either medical or non-medical exemptions, but reported the total number of students with completed vaccinations, the total exemptions was the difference between the number of students enrolled and the number of students completed. This classification process is visualized in Figure 1.

**Figure 1:**
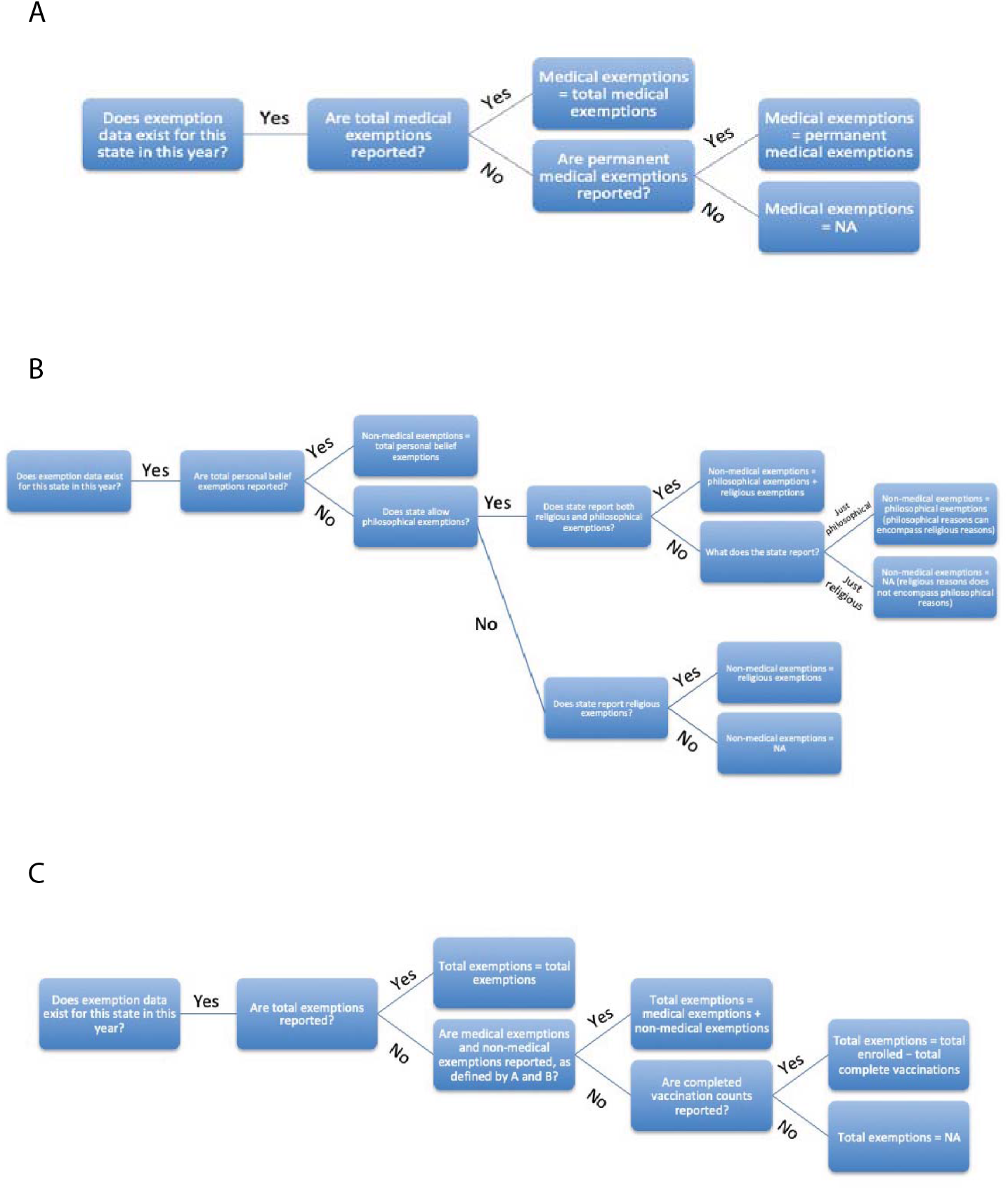
Exemption classification process. Exemptions were classified as medical exemptions (A), non-medical exemptions (B), and total exemptions (C) to standardize reporting across states with different values reported.

We also considered disease-specific exemptions reports. If a state reported the number of exemptions for a vaccine specific to a given infection, that value was used. If the state did not report exemptions, but did provide the total number complete for that disease, the difference between the enrolled students and the completed students was used. For pertussis-specific vaccination, we used DTaP exemptions where available, and TDaP exemptions where DTaP was not available. For measles-specific vaccination, if separate reports were available for measles, mumps, and rubella, the value for measles was used. If measles was not available, then the mumps or rubella exemptions were used, if available.

The data in the figures is only data reported for kindergartens in states where kindergarten-specific data was available, or K-12 data in states where kindergarten-specific data was not reported. States reported age groups heterogeneously, and data by other age groups is available in the data file.

### Code availability

The code used to produce the figures included in the manuscript as well as the full cleaned and raw datasets are available on Github at https://github.com/bansallab/exemptions-landscape. The code runs in Python 3.6.

### Data Records

A master file containing cleaned and consistent data is available on Dryad [15]. This dataset contains the year, state, county FIPS code, and pertinent age group for each record. Each county-year combination that is available has entries for total enrolled, total medical exemptions, total non-medical exemption, religious exemptions, philosophical exemptions, pertussis exemptions, measles exemptions, varicella exemptions, and flu exemptions. State-year available is detailed in Table 1. Additional raw data files are also available.

### Technical Validation

Figure 2 represents the number of total exemptions at the county level reported in 2017. States in blue are those that report kindergarten specific data. States in green report data for kindergarten through 12th grade. The values reported are the total number of exemptions divided by the total number of enrolled students. The highest values, represented in the lightest green or blue colors, represent the counties in which herd immunity appears to be below the 95% threshold for measles. Some states exhibit high levels of heterogeneity in exemption values, like Florida and Pennsylvania. Several states have a number of counties with low exemption rates, like New York and California, as this data was reported after California disallowed non-medical exemptions. More concerning is the fact that several states appear to have many counties with high levels of exemptions, like Washington and Idaho. We also highlight the vast number of states in white which do not report fine-scale exemption data. Increased reporting will be crucial to further understanding the landscape of immunity in the United States.

**Figure 2:**
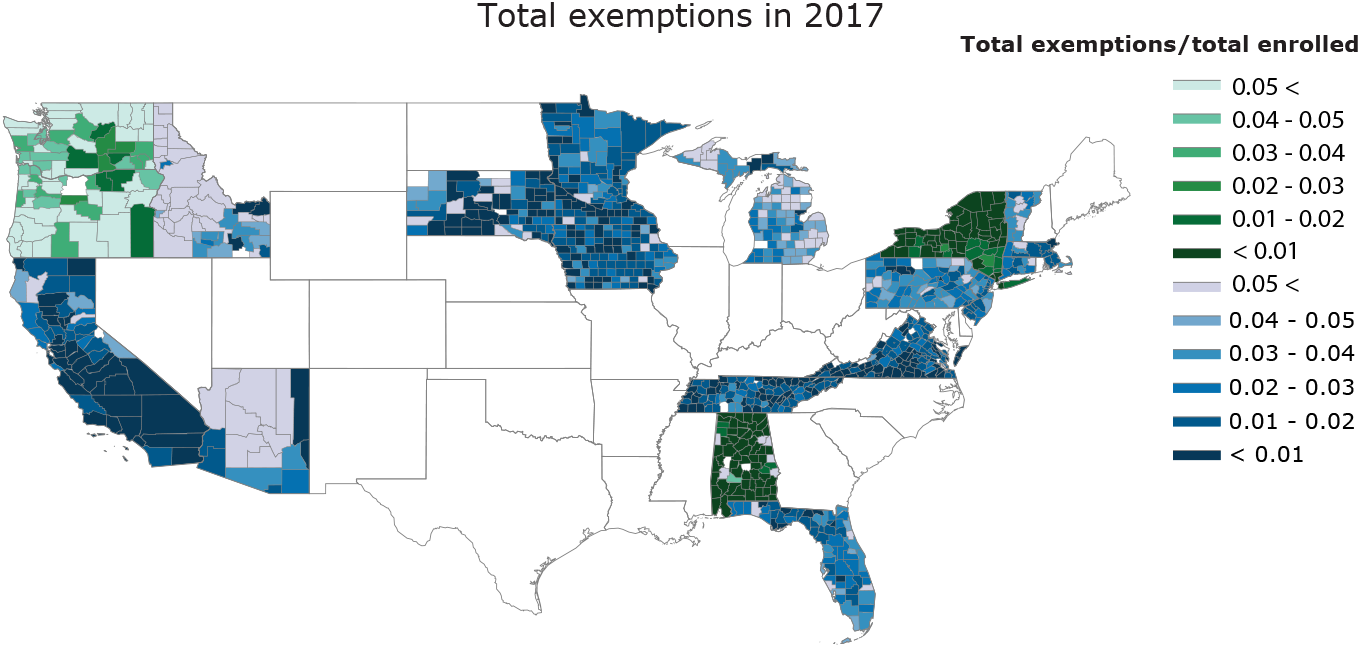
Vaccine exemptions rates are heterogeneous across the United States at a fine spatial scale. Total vaccine exemption rates are reported at the county-level in 2017. Blue-hued states indicate Kindergarten specific data. Green-hued states indicate Kindergarten-12th grade reported data. White indicates missing data.

After demonstrating the spatial distribution of exemptions, we considered the temporal variation in exemptions. Figure 3 shows the time series of total exemptions by state with standard error. States are visible on the plot only in the years in which data was available for more than one year. States are split into different plots only to aid with visual differentiation. This figure highlights the heterogeneity in temporal reporting across states which adds to the difficulty in understanding trends in exemptions and how they might relate to trends in policy changes, vaccination dynamics, or disease dynamics. We also generally see an increase in exemptions over time in the majority of states. Increasing levels of exemptions may reduce herd immunity levels, and this is a problem across the United States. The case of California highlights the importance of studying exemption trends across time. We see that there is a plunge in exemptions following the removal of non-medical exemptions followed by an increase in the exemption level, perhaps indicating that vaccine hesitant individuals found other means to obtain exemptions [4, 16, 3].

**Figure 3:**
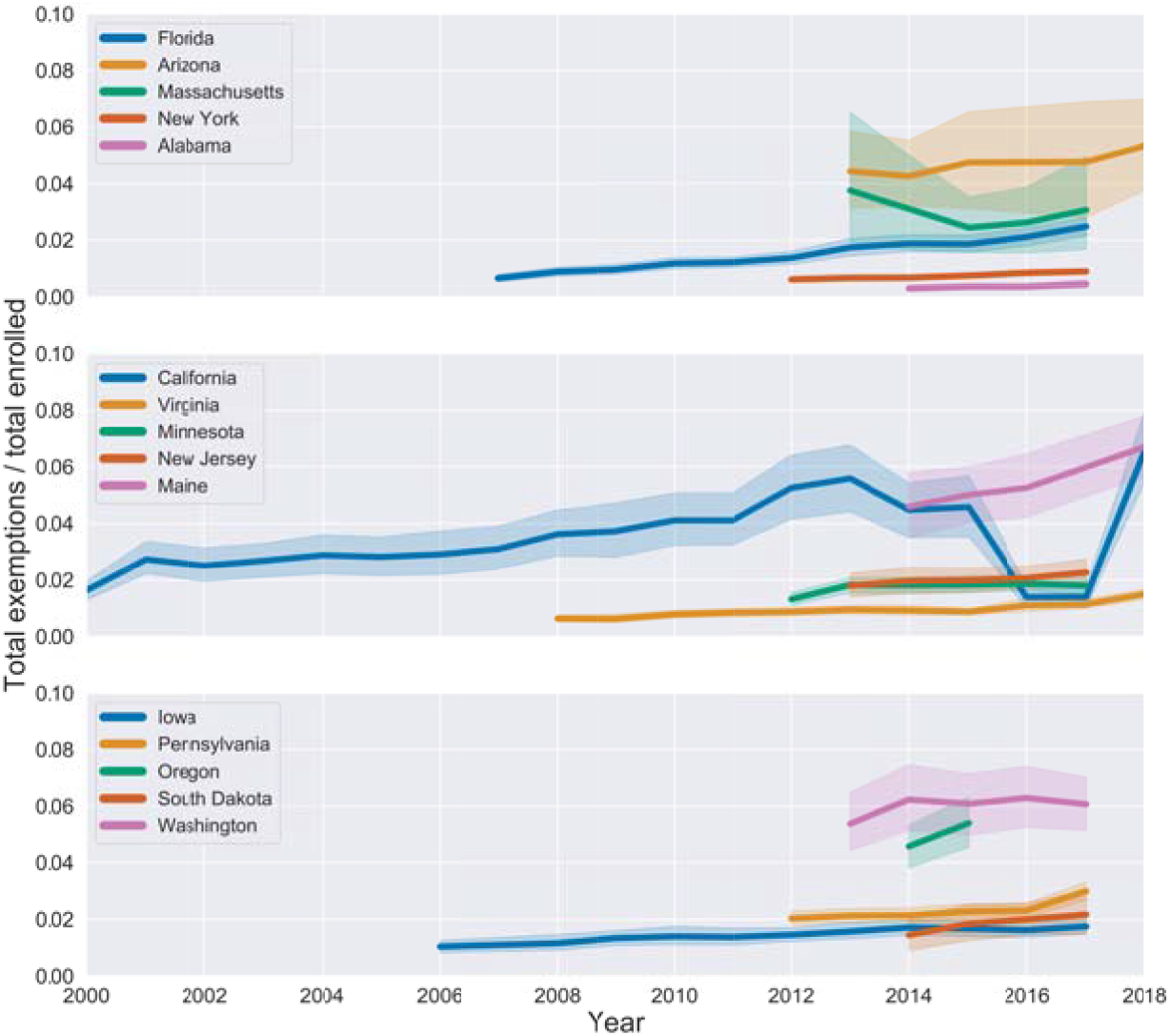
Vaccine exemptions are increasing over time in the majority of states. Total exemption rates are reported at the state-level for each year in which the data was available for more than 1 year. States are located on different plots for visual purposes only.

Figure 4 demonstrates total exemptions across data reported years by state, broken down into medical exemptions, religious exemptions, philosophical exemptions, or unknown. In states that allow philosophical exemptions, these types of exemptions make up the majority of exemptions for most states (Maine, Michigan, and Washington), except for Pennsylvania. Overall, medical exemptions compose a small proportion of total exemptions. In some states, disease-specific exemptions were also reported, and the total exemptions in reported years are shown by state (Figure 5). In several states, like Arizona, Connecticut, Idaho, Minnesota, Vermont, and Washington, it appears that exemptions are consistent for disease types; possibly indicating that children are exempt from all vaccines, instead of obtaining exemptions for specific vaccines. In other states, exemptions for a specific vaccine or vaccines appear greater. Maine and Pennsylvania have more exemptions for varicella. California, Oregon, and Massachusetts have more exemptions for pertussis.

**Figure 4:**
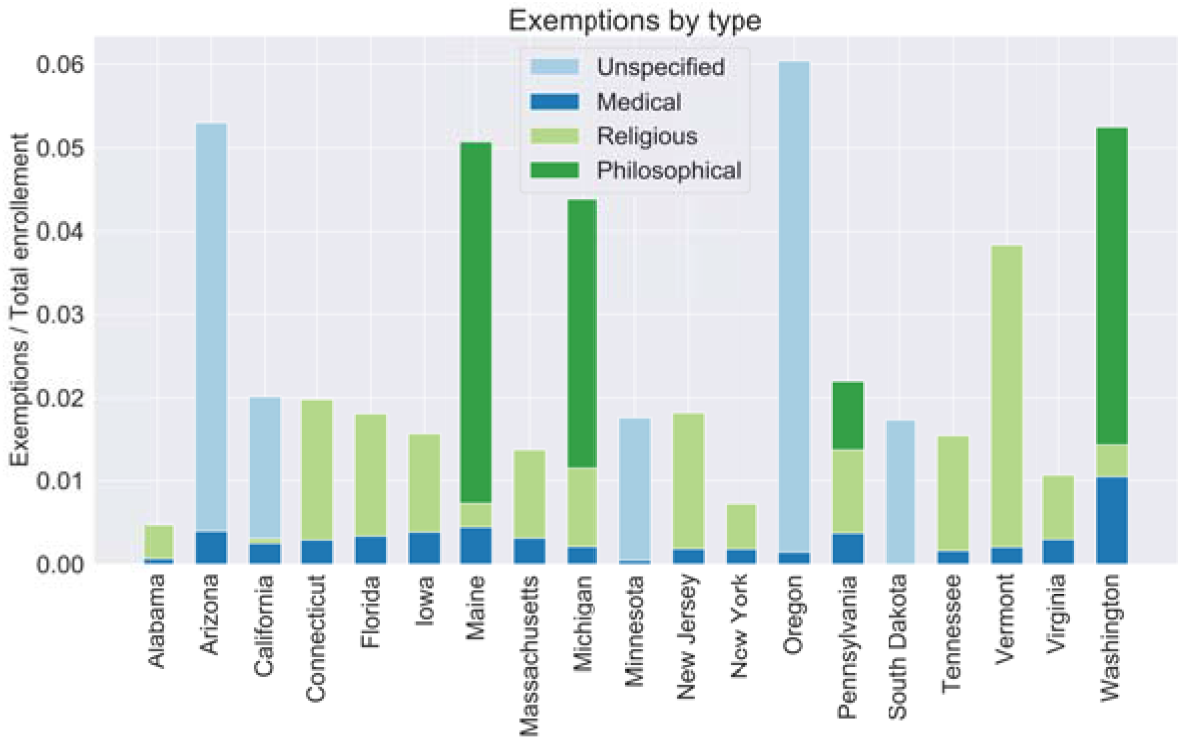
Exemption types depend on the state non-medical exemptions laws, but are dominated by non-medical exemptions. The total of each bar represents the total exemption rate totaled across the available data years for each state. Light blue indicates unspecific exemption types. Dark blue indicates medical exemptions. Green represents non-medical exemptions, where light green shows religious exemptions and dark green shows philosophical exemptions.

**Figure 5:**
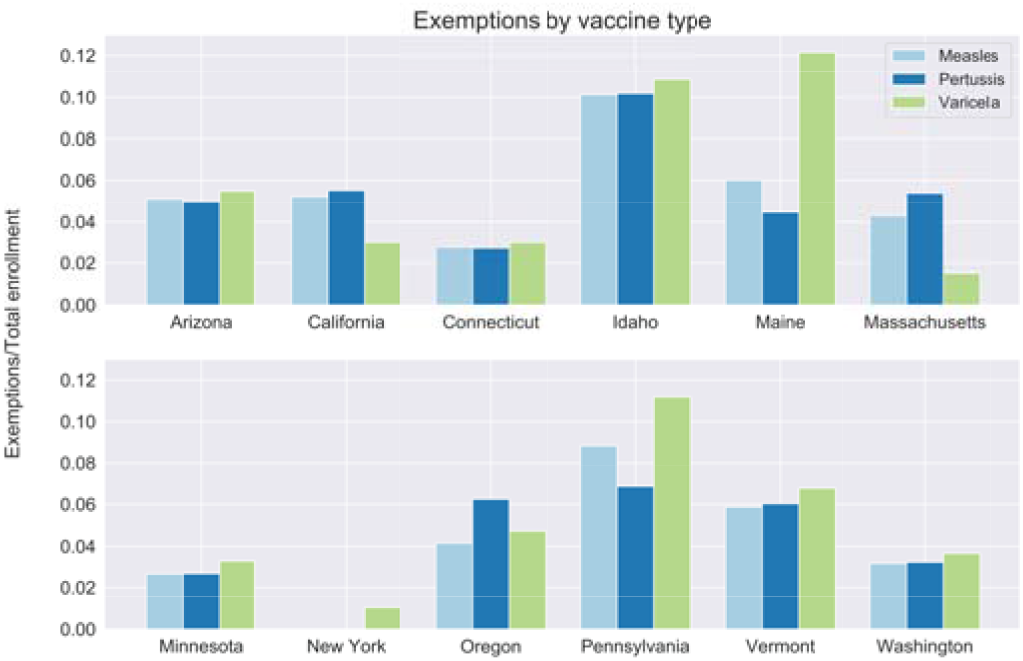
Disease-specific exemption levels vary by state, but are largely consistent across diseases. Measles exemptions (light blue), pertussis exemptions (dark blue), and varicella exemptions (light green) totaled for reported data years are shown for each state that reported disease-specific information.

### Usage Notes

This dataset will have potential uses for guiding public health policy, parameterizing epidemiological models, and validating other sources of vaccination data.

In public health policy, it is important for states to understand the fine-scale landscape of immunity for public health planning. This is necessary for proactive steps, like spatially targeted vaccination campaigns, and for more reactive steps, like anticipating upcoming outbreaks in areas where herd immunity has been waning. Our dataset also makes it feasible to compare across geography and time. A comparative approach is crucial to a better understanding of the underlying drivers of vaccine refusal and the effectiveness of existing public health policies. The larger geographic scope is also crucial to understand the effects of vaccine exemptions at a regional scale; disease susceptibility and transmission does not stop at state borders, thus it’s important for individual states to understand the impact of declining vaccination coverage and herd immunity in neighboring states.

This data is also important for the parameterization of epidemiological models. Incorporating fine-scale population-level immunity into models to further understand and predict the reemergence of childhood diseases in the United State will vastly improve model results. Many prior epidemiological studies have only been able to focus on small spatial scales due to available data [17], but we have seen prior outbreaks of childhood diseases that are not spatially continuous, like the Disneyland measles outbreak [18]. Thus, prediction and mechanistic understanding of childhood disease resurgence will be greatly enhanced by use of this dataset.

Our dataset provides the first unified source of information for the landscape of vaccine refusal for childhood diseases in the United States. School records of immunization provide finer spatial resolution, high response rate, and vaccine-specific information. However, these data are limited in accuracy [19], are only accessible for about half of US states, and remain challenging to compare given differences in data collection methods and data quality [20, 21, 22, 14]. Novel data sources must be explored to provide a truly comparable fine-scale understanding of the heterogeneity in vaccine refusal across all states of the US. With such alternative datasets, validation will be important against our school vaccine exemption dataset.

## Data Availability

All data is available in a Github repository at https://github.com/bansallab/exemptions-landscape

## Acknowledgements

Research reported in this publication was supported by the National Institute Of General Medical Sciences of the National Institutes of Health under Award Number R01GM123007. The content is solely the responsibility of the authors and does not necessarily represent the official views of the National Institutes of Health. We also acknowledge support from the PhRMA Foundation.

## Author contributions

SB conceptualized the work. MK and RG collected and formatted the raw data. CMZ and RG cleaned and analyzed data. CMZ visualized data. CMZ and RG wrote the first draft of the manuscript. CMZ, RG and SB interpreted the findings and edited the manuscript. All authors approved the final manuscript.

